# Scalable, high-purity isolation of blood extracellular vesicles via a cleavable DNA–lipid anchor

**DOI:** 10.1101/2025.09.29.25336888

**Authors:** Stephanie J. Zhang, Rui Fu, Zhen Jiang, Shin-Chin Wang, Michael Lewandowski, Yamuna Krishnan, David R. Walt

## Abstract

Extracellular vesicles (EVs) provide a protected molecular record of disease activity, but their potential as blood-based biomarkers has been limited by isolation methods that are slow, impure, and poorly scalable. We developed a cleavable DNA–lipid anchor (cDLA) that reframes EV isolation as a modular chemistry platform, enabling rapid, economical bead-based capture and release of intact vesicles. In plasma, cDLA achieved markedly higher purity than size-exclusion chromatography, delivering deeper proteomic coverage. We applied the approach to uncover vesicular tau signals that more faithfully tracked with cerebrospinal fluid, improving diagnostic discrimination over bulk plasma tau. cDLA provides a robust and clinically scalable method for EV isolation, opening a path to liquid biopsy in neurodegenerative diseases and beyond.

---

EVs carry molecular cargo that reflects the state of otherwise inaccessible tissues and are widely pursued as blood-based biomarkers.^1, 2^ Isolation of EVs from biofluids remains a major barrier to their use as clinical biomarkers. Existing methods, such as ultracentrifugation and size-exclusion chromatography (SEC), are poorly scalable and yield fractions contaminated with abundant plasma proteins, thereby restricting their use in large clinical studies and proteomic analyses.^3, 4^

To address this challenge, we developed a cDLA strategy that enables rapid magnetic capture and release of intact EVs (**Fig. 1a**). A lipid–DNA probe^5^ pre-annealed to a complementary desthiobiotin (dB)–DNA strand inserts into EV membranes, enabling streptavidin-bead capture and release by DNase digestion or biotin competition. Flow cytometry and fluorescence microscopy confirmed efficient probe insertion and vesicle capture (**Fig. 1b, i-ii**). Control experiments verified that capture required the lipid–DNA anchor and was not due to nonspecific binding (**Extended Data Fig. 1**).

**Fig. 1.**
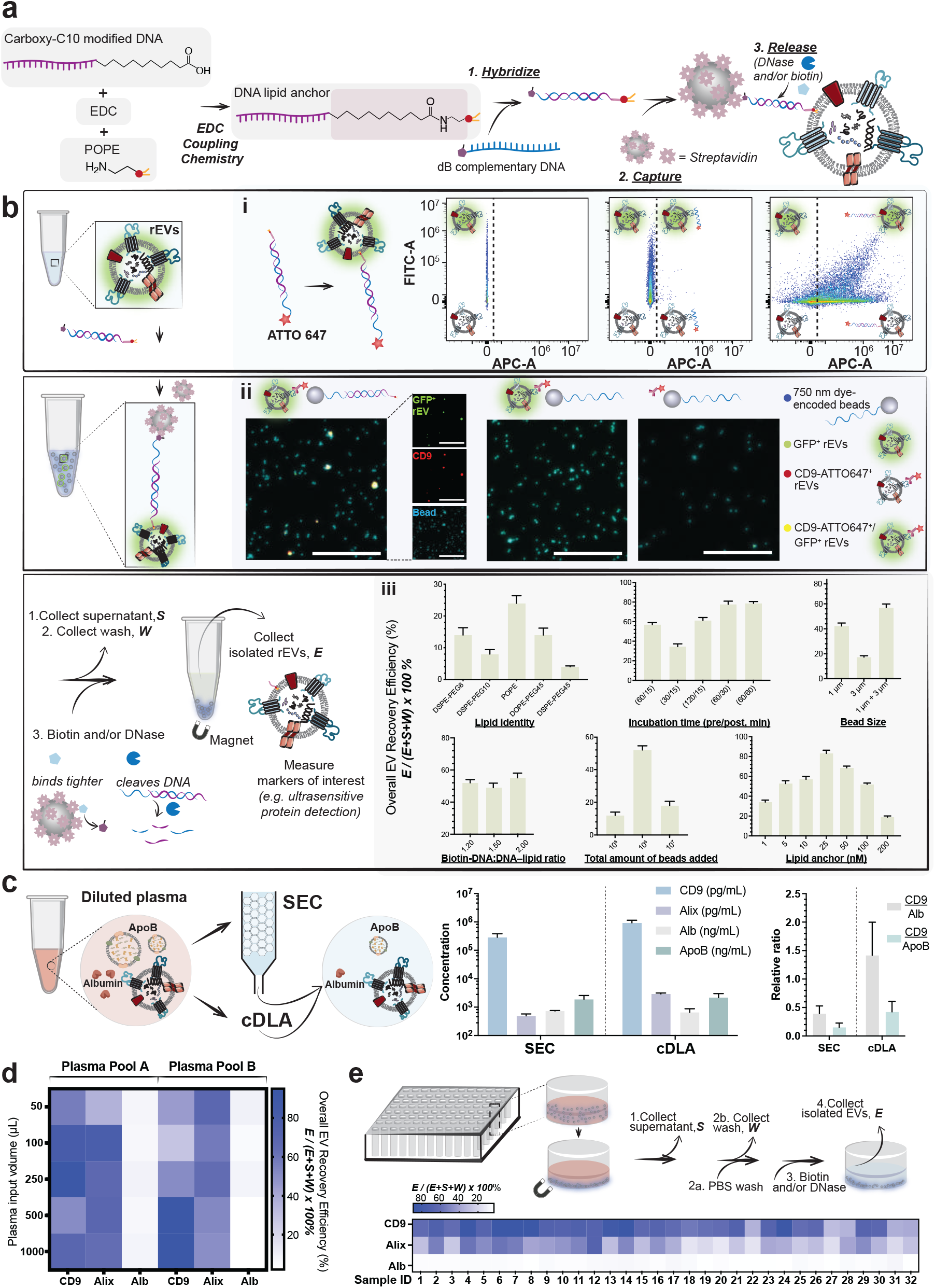
Cleavable DNA–lipid anchor enables scalable and high-purity EV isolation. **a**, Schematic of the cDLA workflow. POPE–DNA was synthesized via EDC coupling (other lipid–DNA conjugations in Methods). The lipid–DNA probe hybridizes with a complementary desthiobiotin–DNA strand, inserts into EV membranes, and the duplex is captured on streptavidin beads. EVs are released by DNase digestion and/or biotin competition. **b**, Workflow and optimization for EV capture and release. (i) Flow cytometry shows efficient rEV (GFP^+/-^) membrane labeling by lipid–DNA, with ATTO647 signal reporting successful insertion. (ii) Fluorescence microscopy shows co-localization of bead-bound rEVs (GFP^+^) with ATTO647 signal; in the left micrograph, separate channels highlight colocalization of GFP^+^ rEV, CD9, and bead signals (quantitative analysis in Fig. **S2**; scale bar, 100 µm). (iii) Recovery efficiency across lipid identity, incubation time, bead size, DNA–lipid/dB–DNA ratio, bead input, and lipid anchor concentration. **c**, EV markers (CD9, Alix) are enriched, while abundant plasma proteins including apolipoprotein B (ApoB) and Alb are depleted; CD9/Alb and CD9/ApoB ratios are increased with cDLA compared to SEC. **d**, Recovery of EV markers versus plasma contaminants across two plasma pools at increasing input volumes (50–1000 µL). **e**, High-throughput implementation in 96-well format showing consistent EV recovery and contaminant depletion across 32 plasma samples.

We first optimized capture conditions in a recombinant EV (rEV) system,^6^ quantifying recovery by normalized detection of CD9,^7^ a canonical EV tetraspanin marker (**Fig. 1b, iii**). Among lipid anchors, POPE yielded the highest recovery (∼24% versus <14% for other lipids; see **Supplementary Text** for discussion of lipid identity). Mixed 1 µm + 3 µm beads outperformed single sizes by ∼15%. Recovery displayed a bell-shaped dependence on bead and probe inputs, with intermediate levels performing best. Longer insertion times improved capture efficiency, increasing from ∼35% at 10 min to ∼57% at 60 min. Under optimized conditions, cDLA recovered up to ∼80% of input rEVs, with intact release by DNase or biotin; further optimization details are shown in **Fig. S1**.

We next tested cDLA in human plasma, where EV recovery is complicated by abundant free proteins such as albumin (Alb) and apolipoproteins.^8^ After tuning parameters (**Extended Data Fig. 2**), cDLA enriched canonical EV markers (CD9, Alix) while minimizing protein carryover (**Fig. 1c**). Vesicle integrity was confirmed by protease protection assays: internal EV proteins (e.g., Alix) were shielded from proteinase K unless detergent was added, whereas external proteins such as Alb were digested by PK alone. Both cDLA and SEC preserved vesicle integrity, but cDLA fractions displayed a higher Alix-to-Alb ratio (**Fig. S3**).

To assess scalability, we applied cDLA across plasma volumes from 50 to 1000 μL. Recovery of CD9 and Alix remained consistent while Alb was strongly excluded (**Fig. 1d**). Adaptation to a 96-well magnetic bead format enabled analysis of 32 individual plasma samples in parallel, with CD9 and Alix recovery consistently exceeding 60–70% and Alb strongly depleted, demonstrating robustness across samples and compatibility with high-throughput workflows (**Fig. 1e**).

We then benchmarked cDLA against SEC by profiling plasma EV isolates with mass spectrometry (**Fig. 2a**). EVs isolated by cDLA yielded ∼6-fold greater proteome depth, with 80% of identified proteins overlapping with ExoCarta entries^9^ compared with 70% for SEC (**Fig. 2b**). This translated to a substantially higher proportion of EV-associated proteins unique to the cDLA isolates, while SEC contributed only a small minority. Within the canonical panel of 12 EV markers, EVs isolated by cDLA contained nine, whereas SEC isolates contained only one (**Fig. 2c**). Importantly, contaminants such as albumin and apolipoproteins comprised a far larger share of the proteomic signal in SEC than in cDLA isolates (**Fig. 2d**), indicating that contaminants likely mask detection of lower-abundance vesicular proteins. The enhanced depth achieved with cDLA enabled detection of neuron-associated proteins (e.g., tau, TDP43, α-synuclein)^10, 11^ and endothelial markers (e.g., PECAM1),^12^ which are increasingly recognized as clinically relevant EV biomarkers. EVs isolated by cDLA therefore more accurately represent the EV proteome by enriching bona fide vesicular proteins while reducing interference from abundant plasma proteins.

**Fig. 2.**
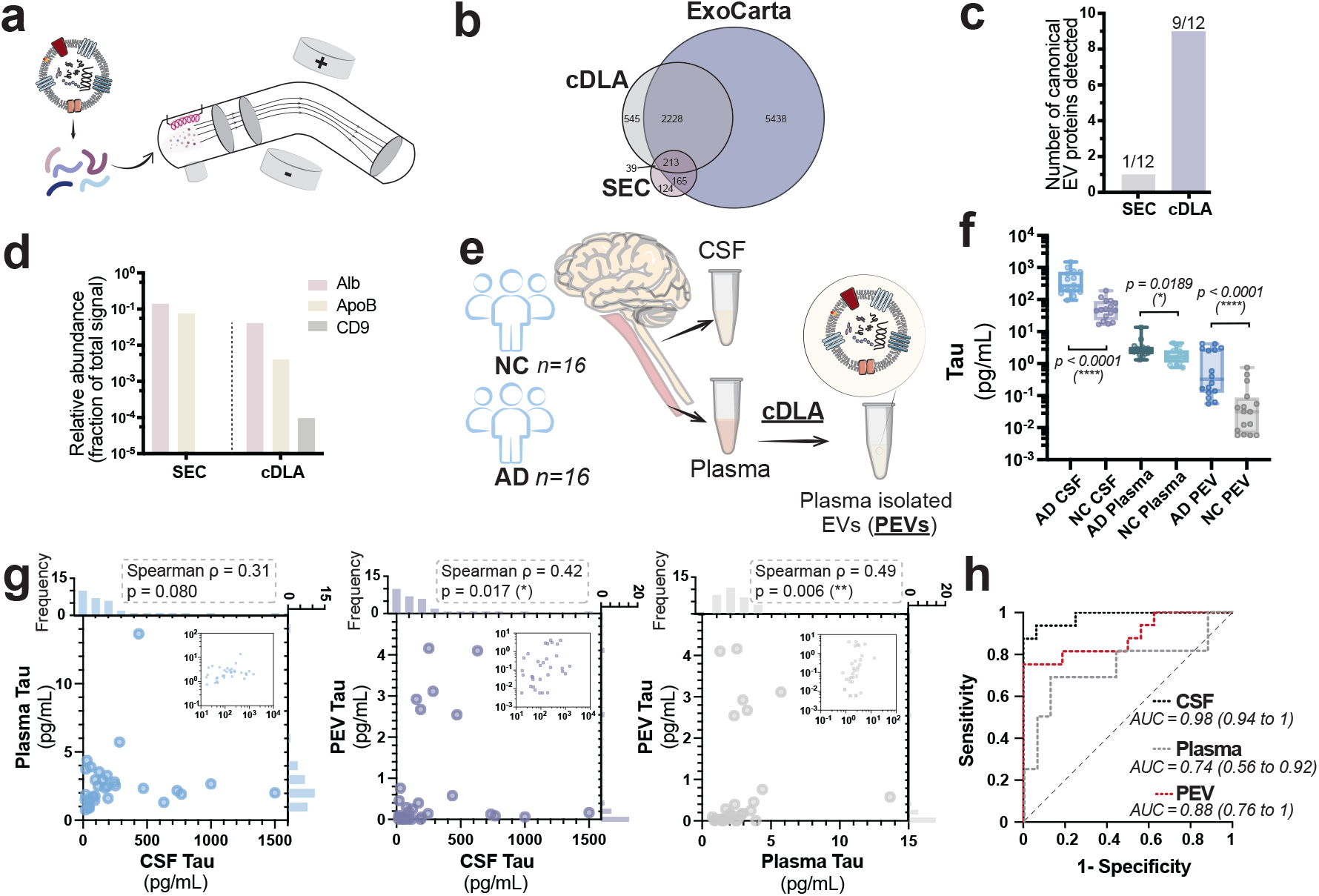
High-purity EV isolation by cDLA enables downstream biomarker applications. **a**, Schematic of mass spectrometry analysis of EVs isolated by cDLA and SEC and **b**, proteomic comparison with ExoCarta mass spectrometry entries. **c**, Nine of twelve canonical EV proteins (CD9, CD63, CD81, TSG101, Alix/PDCD6IP, HSPA1A, HSPA8, HSP90AA1, FLOT1, FLOT2, ANXA2, ANXA5) were detected in cDLA isolates versus one in SEC. **d**, Plasma contaminants were reduced in cDLA isolates, with Alb (P02768, P02769) and ApoB (P04114) comprising a smaller fraction of the proteomic signal compared to SEC, while CD9 was detectable in cDLA isolates but absent in SEC. **e**, Paired CSF and plasma from AD (n=16) and NC (n=16) were analyzed; PEVs were isolated using cDLA method. **f**, Tau was significantly elevated in AD versus NC for CSF and PEVs (p<0.0001), with plasma showing a modest difference (p=0.0189). **g**, PEV tau correlated more strongly with CSF tau (ρ=0.42, p=0.017) than with plasma tau (ρ=0.31, p=0.080), and also correlated with plasma tau (ρ=0.49, p=0.006). Insets show the same data on a log–log scale. **h**, ROC curves show that PEV tau improved discrimination of AD from NC (AUC=0.88) compared to plasma tau (AUC=0.74), though CSF tau remained highest (AUC=0.98).

Having established the robustness of cDLA, we next asked whether the platform could unlock clinically relevant signals. Plasma tau has long been explored as a blood-based biomarker, yet its concentrations are low, variable, and weakly correlated with CSF,^13^ in part because circulating tau includes substantial contributions from peripheral tissues. EVs can cross the blood–brain barrier, raising the possibility that tau within plasma EVs may better capture CNS input. While encapsulation could protect tau from any source, neuronal secretion of tau in EVs is well established,^10, 14^ whereas peripheral sources remain less clear. This provided the rationale to test whether cDLA-isolated plasma EV tau (PEV tau) more closely reflects CSF tau than bulk plasma tau.

Using cDLA—which isolates 32 plasma samples in under three hours and scales seamlessly to 96 or more samples—we coupled rapid EV purification with ultrasensitive digital protein detection (**Fig. 2e**).^15, 16^ PEV–associated tau was quantitatively less abundant than total plasma tau (**Fig. 2f**), consistent with EVs representing a sub-fraction of circulating tau. Despite lower concentrations, PEV tau was significantly elevated in Alzheimer’s disease (AD) compared to controls and tracked more closely with CSF tau (ρ=0.42, p=0.017) than plasma tau (ρ=0.31, p=0.080) (**Fig. 2g**). PEV tau also correlated with plasma tau (ρ=0.49, p=0.006), suggesting overlapping but distinct sources.

This selective enrichment improved diagnostic discrimination for AD: PEV tau achieved an AUC of 0.88 versus 0.74 for plasma tau, approaching CSF tau (AUC 0.98) (**Fig. 2h**). Thus, plasma EV tau provides a more informative signal for disease detection and tracks more closely with CSF tau than bulk plasma tau. PEV tau correlated with both plasma and CSF tau, suggesting contributions from overlapping sources but also reflecting distinct information not captured by either alone (**Fig. 2g)**. While we cannot definitively ascribe the improvement to selective brain enrichment, the stronger diagnostic performance suggests that EV isolation enhances access to clinically relevant signals, including but not limited to CNS contributions. More broadly, this principle may extend beyond tau, as EVs can stabilize otherwise labile proteins in circulation, protecting them from degradation and enabling molecular readouts inaccessible from bulk plasma.

cDLA reframes EV isolation as an engineered chemistry platform rather than a fractionation problem. Conventional methods such as ultracentrifugation or SEC depend on bulk physical separations with an inevitable trade-off between purity, throughput, and vesicle integrity. In contrast, cDLA integrates lipid chemistry, nucleic acid modularity, and magnetic capture to recover intact EVs at scale. This modular design also enables multiplexing: DNA-coded beads capture complementary DNA–tagged antibodies or aptamers, assigning EV subsets to defined bead codes in a single incubation. Thus, cDLA can recover rare, disease-relevant vesicles from a background of abundant carriers.

From a translational perspective, cDLA enables high-purity EV isolation at a cost of <$1 per sample and can be readily integrated into multiwell magnetic bead workflows or robotic platforms. This scalability reduces trial-scale isolation from days to hours and makes systematic, large-cohort analyses of plasma EVs feasible. By preserving intact vesicles for downstream multiomic readouts, cDLA not only improves biomarker detection in AD but also establishes a general framework for accessing molecular information in blood that was previously obscured by plasma proteins. By uniting modular chemistry with clinical scalability, it transforms EV isolation from a methodological bottleneck into a practical foundation for next-generation liquid biopsies.

## Data Availability

All data produced in the present study are available upon reasonable request to the corresponding authors.

## References

1. Shah, R. et al. Circulating Extracellular Vesicles in Human Disease. New Engl J Med 379, 958–966 (2018).

2. Tkach, M. et al. Communication by Extracellular Vesicles: Where We Are and Where We Need to Go. Cell164, 1226–1232 (2016).

3. Kumar, M.A. et al. Extracellular vesicles as tools and targets in therapy for diseases. Signal Transduct Tar 9(2024).

4. Allelein, S. et al. Potential and challenges of specifically isolating extracellular vesicles from heterogeneous populations. Sci Rep-Uk 11 (2021).

5. Saminathan, A. et al. A DNA-based voltmeter for organelles. Nat Nanotechnol 16, 96–103 (2021).

6. Geeurickx, E. et al. The generation and use of recombinant extracellular vesicles as biological reference material. Nat Commun 10 (2019).

7. Norman, M. et al. L1CAM is not associated with extracellular vesicles in human cerebrospinal fluid or plasma. Nat Methods 18, 631–634 (2021).

8. Sódar, B.W. et al. Low-density lipoprotein mimics blood plasma-derived exosomes and microvesicles during isolation and detection. Sci Rep-Uk 6 (2016).

9. Keerthikumar, S. et al. ExoCarta: A Web-Based Compendium of Exosomal Cargo. J Mol Biol 428, 688–692 (2016).

10. Chatterjee, M. et al. Plasma extracellular vesicle tau and TDP-43 as diagnostic biomarkers in FTD and ALS. Nat Med 30, 1771–1783 (2024).

11. Gilboa, T. et al. Measurement of α-synuclein as protein cargo in plasma extracellular vesicles. P Natl Acad Sci USA 122, e2408949121 (2024).

12. Mazzucco, M. et al. CNS endothelial derived extracellular vesicles are biomarkers of active disease in multiple sclerosis. Fluids Barriers Cns 19 (2022).

13. Fossati, S. et al. Plasma tau complements CSF tau and P-tau in the diagnosis of Alzheimer’s disease. Alzheimers Dement (Amst) 11, 483–492 (2019).

14. Saman, S. et al. Exosome-associated Tau Is Secreted in Tauopathy Models and Is Selectively Phosphorylated in Cerebrospinal Fluid in Early Alzheimer Disease. J Biol Chem 287, 3842–3849 (2012).

15. Zhang, S.J. et al. A Multiplexed Digital Platform Enables Detection of Attomolar Protein Levels with Minimal Cross-Reactivity. Acs Nano 18, 29891–29901 (2024).

16. Rissin, D.M. et al. Single-molecule enzyme-linked immunosorbent assay detects serum proteins at subfemtomolar concentrations. Nat Biotechnol 28, 595–599 (2010).

