## Supplementary Information for "Scalable, high-purity isolation of blood extracellular vesicles via a cleavable DNA–lipid anchor"

### Supporting Information
#### Materials and Methods
##### Materials
All affinity reagents, recombinant proteins, and DNA oligonucleotides used in this work are listed in the Supporting Information (**Tables S1-3**). Buffers were from Quanterix and Invitrogen (Thermo Fisher Scientific); paramagnetic beads from Quanterix, New England Biolabs, and Bangs Laboratories; lipids from Avanti Polar Lipids; and conjugation reagents from Thermo Fisher Scientific and Millipore Sigma. Sources of additional reagents are provided at first mention.

##### Synthesis and characterization of lipid–DNA conjugates
POPE–DNA conjugates were synthesized using a 5′-carboxyl-C10–modified DNA strand (Integrated DNA Technologies; sequence in **Table S1**). POPE (Avanti Polar Lipids) was coupled to the carboxyl-modified oligonucleotide via amide bond formation, and the conjugates were purified and lyophilized. The product identity was confirmed by denaturing PAGE.

Other lipid–DNA conjugates were prepared following established DBCO–azide (Thermo Fisher Scientific) coupling protocols,[1] substituting different lipid–azide precursors as required. Briefly, a 5′-DBCO–modified 24-nt DNA strand (20 µM; Integrated DNA Technologies; sequence in **Table S1**) was reacted with lipid–azide (40 µM, 2 eq.) in 20 mM sodium phosphate buffer (pH 7.4) for 4 h at room temperature. Final products were characterized by mass spectrometry, which confirmed the expected molecular masses (**Fig. S4**).

##### EV isolation by cDLA
DNA–lipid conjugates were annealed with complementary DNA strands at final concentrations of 1 µM (lipid–DNA) and 1.2 µM (complementary strand; sequence provided in **Table S1**) in hybridization buffer containing 10 mM Tris (Invitrogen, Thermo Fisher Scientific; pH 8.0) and 100 mM NaCl (Invitrogen, Thermo Fisher Scientific). Annealing was carried out by heating samples to 95 °C for 5 min, followed by controlled cooling to 23 °C at 0.1 °C/s using a thermocycler (Bio-Rad).
For capture experiments, the pre-annealed DNA duplex mixture was added to diluted plasma samples or rEVs (Sigma-Aldrich). Samples were incubated for 1 h at room temperature with gentle orbital agitation (100 rpm) to allow incorporation of DNA–lipid conjugates into EV membranes. Streptavidin-coated magnetic beads were then introduced (plasma diluted 5-fold in 1x PBS; Gibco, Thermo Fisher Scientific). Two bead formulations were tested: (i) 1 µm diameter streptavidin-coated beads (New England Biolabs) and (ii) 3 µm diameter streptavidin-coated beads (Bangs Laboratories, ProMag 3 series). After bead addition, suspensions were mixed intermittently at 120 rpm for 10 s every 10 min during a 1 h incubation at room temperature.
Following incubation, supernatants were collected, and bead pellets were washed once with 1x PBS; the first wash fractions were retained separately. Beads were then washed a second time, transferred into fresh low-retention tubes, and the second wash fractions were also collected. Beads were subsequently resuspended in DNase reaction mixture, in biotin solution, or sequentially in DNase followed by biotin, depending on the downstream application. Initial lipid and bead concentrations, as well as the incubation times, were based on preliminary estimates described in the **Supplementary Text B-D**.
To enzymatically release EVs from DNA tethers, nuclease solution was added to bead suspensions, followed by incubation at 37 °C with orbital shaking at 650 rpm for 30 min. EVs were then released either sequentially—by Benzonase treatment followed by addition of biotin (Sigma-Aldrich; mixed for 5 min at room temperature)—or directly by biotin addition alone. In both cases, biotin saturated residual streptavidin sites and prevented re-binding of DNA strands. For samples intended for downstream mass spectrometry, only the biotin-mediated release was performed without Benzonase digestion.

##### Flow cytometry for detection of DNA–lipid–tagged EVs
DNA–lipid conjugates were annealed with complementary DNA strands labeled with ATTO647 at final concentrations of 1 µM (lipid–DNA) and 1.2 µM (complementary strand) in hybridization buffer (10 mM Tris, pH 7.5, 100 mM NaCl). Annealing was performed by heating to 95 °C for 5 min, followed by controlled cooling to 23 °C at 0.1 °C/s in a thermocycler.
For flow cytometry experiments, the pre-annealed DNA duplex mixture was added to rEVs. Suspensions were gently mixed by inversion (three times) and incubated at room temperature for 15 min to allow incorporation of DNA–lipid conjugates into rEV membranes. Samples were then diluted 5-fold in 1x PBS to a final volume of 50 µL. Flow cytometry was performed using a NovoCyte flow cytometer (Agilent Technologies). ATTO647–

labeled DNA was detected in the APC channel (emission filter centered at 660 nm). A minimum of 100 µL was acquired per sample.

**Fluorescence microscopy to confirm DNA–lipid insertion and bead capture**

DNA was conjugated to 750-nm dye–encoded beads as previously described.[2] To allow membrane insertion, DNA–lipid conjugates were combined with rEVs and incubated for 15 min at room temperature. The resulting complexes were introduced to DNA-coupled beads. Bead-bound EVs were labeled with CD9–ATTO647 antibody (10 min, room temperature). Beads were washed twice with buffer (50 mM Tris-HCl, 50 mM NaCl), concentrated to 10–15 µL, and resuspended.

Suspensions were drop-cast onto microscope slides and allowed to adhere before imaging. Images were acquired on an Olympus IX81 inverted microscope equipped with a 20 x LMPlanFl objective (NA 0.40) and a scientific CMOS camera (ORCA-Flash4.0 LT+, Hamamatsu), with stage and camera control performed using cellSens software (Olympus). Fluorescence channels were collected using a GFP filter (1 s exposure, rEVs), Cy5 filter (5 s exposure, CD9–ATTO647), and Cy7 filter (1 s exposure, dye-encoded beads); brightfield and fluorescence channels were recorded for each frame. Post-acquisition analysis was performed using ThunderSTORM (ImageJ plugin) for spot selection.

**Preparation of capture and labeling reagents**

Capture antibodies (**Table S2**) were buffer exchanged with Bead Conjugation Buffer (Quanterix) using a 50K Amicon Ultra-0.5 mL centrifugal filter (Millipore Sigma). Antibody solutions were brought to 500 µL with Bead Conjugation Buffer and centrifuged three times at 14,000 x g for 5 min, with 450 µL Bead Conjugation Buffer added between cycles. Buffer-exchanged antibodies were recovered by inverting the filter into a new tube and centrifuging at 1,000 x g for 2 min, followed by rinsing the filter with 50 µL Bead Conjugation Buffer and centrifuging again at 1,000 x g for 2 min. Antibody concentration was measured using a NanoDrop spectrophotometer.

For each bead type, the indicated number of beads (**Table S4**) was washed three times with 300 µL Bead Wash Buffer (Quanterix) and twice with 300 µL Bead Conjugation Buffer, then resuspended in cold Bead Conjugation Buffer. A 1 mg vial of 1-ethyl-3-(3-dimethylaminopropyl)carbodiimide hydrochloride (EDC; Thermo Fisher Scientific) was dissolved in 100 µL cold Bead Conjugation Buffer, and the desired volume was added to the beads. Beads were shaken for 30 min at either room temperature or 4 °C. After EDC activation of bead carboxyl groups, beads were washed once with 300 µL cold Bead Conjugation Buffer and resuspended in the buffer-exchanged antibody solution. Antibody conjugation was performed by shaking the beads for 2 h at either room temperature or 4 °C, followed by washing twice with 300 µL Bead Wash Buffer.

The antibody-coupled beads were blocked for 30 min at room temperature with shaking in 300 µL Bead Blocking Buffer (Quanterix). After washing once each with 300 µL Bead Wash Buffer and Bead Diluent (Quanterix), beads were resuspended in 200 µL Bead Diluent, counted using a Beckman Coulter Z1 Particle Counter, and stored at 4 °C. Detector antibodies for non-barcoded MOSAIC assays were obtained in biotinylated form.

**Preparation of antibody-DNA conjugates**

*Circularized padlock DNA template-primer hybrid.* For each detector antibody, a 5′ azide-modified primer was annealed to a unique DNA template (**Table S3**) by heating a solution of 30 µM primer and 30.3 µM template in NEBNext Quick Ligation Buffer (New England Biolabs) at 95 °C for 2 min, followed by cooling to room temperature over 90 min. Ligation was then performed by adding T4 DNA ligase and incubating at room temperature for 2 h. The ligation reaction was buffer-exchanged into phosphate-buffered saline (PBS) with 1 mM EDTA using 7K MWCO Zeba spin desalting columns (Thermo Fisher Scientific).

*DBCO-modified antibody.* For conjugation, the detector antibody was either reconstituted into 1x PBS from lyophilized form or buffer exchanged into 1x PBS using a 50K Amicon Ultra-0.5 mL centrifugal filter, incubated with a 20-fold molar excess of dibenzocyclooctyne-PEG4-N-hydroxysuccinimidyl ester (DBCO-PEG4-NHS, Millipore Sigma) for 30 min at room temperature, and purified with a 50K Amicon Ultra-0.5 mL centrifugal filter in 1x PBS with 1 mM EDTA.

*Conjugation of azide-modified DNA to DBCO-modified antibody via copper free click chemistry.* A two-fold molar excess of the ligated primer-template was then added to the DBCO-modified antibody and incubated overnight at 4°C. The conjugate was stored in aliquots at -80 °C in 1x PBS with 5 mM EDTA, 0.1% BSA, and 0.02% sodium azide.

**SIMOA, MOSAIC, and barcoded MOSAIC assays**
All assays in this study were performed in single-plex format using either the Simoa HD-X Analyzer (Quanterix), the MOSAIC platform, or the barcoded MOSAIC platform. Targets included CD9, CD81, albumin (Alb), Alix, tau, and ApoB, with antibody pairs and protein standards listed in **Table S2**. Each assay was validated by spike-and-recovery in biofluids, with acceptable recovery defined as 70–130% (**Table S6**). Calibration curves were used to establish the lower limit of detection (LOD) and lower limit of quantification (LLOQ) for each analyte (**Fig. S5 and Table S7**).
For tau measurements, commercial Simoa kits (Neuro 4 Plex B kit and Tau 2.0 kit; Quanterix) were used to analyze CSF at 10x dilution and plasma at 4x dilution. EV-associated tau was quantified by barcoded MOSAIC assays at 6x dilution. The concordance between Simoa and MOSAIC outputs has been established previously.[3] All samples were assayed in duplicate. Signal outputs were reported as Average Enzyme per Bead (AEB) for Simoa assays (calculated by HD-X software) or as Average Molecule per Bead (AMB) for MOSAIC assays (calculated from flow-based bead counts).

**EV Isolation by SEC**
EVs were isolated from plasma using SEC as previously described.[4] Briefly, Sepharose CL-6B resin was washed three times with an equal volume of 1x PBS in a sterile glass container. An additional equal volume of 1x PBS was mixed with the resin, and the slurry was packed into empty Econo-Pac chromatography columns (Bio-Rad) until a settled bed volume of 10 mL was achieved. A frit was then placed on top of the resin to secure the column. Columns were equilibrated with four successive washes of 5 mL 1x PBS each. Once the final wash buffer had completely drained, 0.25 mL of plasma, pre-diluted to a total volume of 1 mL with 1x PBS, was applied to the column. After the plasma fully entered the resin bed, 1x PBS was added to elute 0.5 mL fractions sequentially. Fractions 1–2, corresponding to the sample entry, were discarded. Fractions 3–6, representing the column void volume, were also discarded. EVs were collected beginning with fraction 7. For experiments requiring total EV recovery, an additional 2 mL of 1x PBS was applied to the column, and fractions 7–10 were collected and pooled.

**Protease protection assay**
Protease protection assays were performed on EVs isolated by either SEC or cDLA capture. SEC-isolated EV preparations were divided into three 200 µL aliquots corresponding to no treatment (NT), protease only (PK), and protease plus detergent (PK+Triton, PKTx) conditions. cDLA-isolated EVs, obtained at higher concentration, were diluted in 1x PBS to match the SEC EV sample volume and similarly divided into three 200 µL aliquots. Proteinase K (PK; Thermo Fisher Scientific) was added to the PK and PKTx groups at a final concentration of 0.2 mg/mL, while an equivalent volume of 1x PBS was added to the NT group. To permeabilize EV membranes and allow protease access to luminal proteins, Triton X-100 (Sigma-Aldrich) was added to the PKTx group at a final concentration of 0.5% (v/v). All samples were incubated at 37 °C for 30 min. Protease activity was quenched by addition of phenylmethylsulfonyl fluoride (PMSF; Roche) to a final concentration of 2.5 mM, followed by shaking at 300 rpm for 1 h at room temperature. To standardize detergent exposure across conditions, Triton X-100 (0.5% v/v) was added post-quenching to both the NT and PK groups.

**Mass spectrometric analysis of EV proteins**
EVs isolated from pooled human plasma by either cDLA or SEC were processed with equal starting amounts of total protein (~2 µg) for each method. Samples were prepared using the Prototype MPSP Mass Spec Sample Prep Kit (Promega). EV aliquots were combined with lysis buffer, mixed briefly, and incubated for 5 min at room temperature. Reduction was performed by adding TCEP (Thermo Fisher Scientific) to a final concentration of 2 mM, followed by incubation for 45 min at room temperature with shaking. Alkylation was carried out with iodoacetamide (Sigma-Aldrich) to a final concentration of 1.5 mM, followed by incubation for 30 min at room temperature with shaking.
Proteins were captured on magnetic beads by adding isopropanol to each sample and shaking for 20 min at room temperature. Beads were isolated on a magnetic rack, supernatants discarded, and washed three times with 80% ethanol. On-bead digestion was performed in digestion buffer containing trypsin at 37 °C overnight with shaking at 1200 rpm. Following digestion, peptide-containing supernatants were recovered by magnetic separation and transferred to fresh low-retention tubes. Samples were concentrated in a SpeedVac (Eppendorf) at 30 °C for 30 min, during which peptide solutions condensed into oil-like droplets, and then resuspended in 0.1% formic acid (Sigma-Aldrich) to an estimated concentration of ~25 ng/µL, based on the

initial protein input. LC–MS/MS analysis was performed on a Thermo Scientific Astral mass spectrometer. Peptides were separated by reverse-phase liquid chromatography using a 25-min gradient and analyzed in data-independent acquisition (DIA) mode. Raw spectra were processed and searched against the UniProt human proteome database using Proteome Discoverer software (Thermo Fisher Scientific), and peptide-spectrum matches were filtered at a 1% false discovery rate (FDR) at both peptide and protein levels.

**Human sample handling**
Plasma and CSF samples from AD patients and NC were obtained from the Massachusetts General Hospital (MGH) Biobank under approved institutional protocols. Plasma and CSF aliquots were thawed at room temperature and centrifuged at 2,000 x g for 10 min to remove cells and large debris. Plasma supernatants were further clarified by centrifugation through 0.45 μm Spin-X centrifuge tube filters (Corning Costar; Sigma-Aldrich) at 2,000 x g for 10 min to eliminate residual cellular material.
For experiments directly comparing EV isolation by SEC and cDLA capture, equal plasma volumes from multiple individuals were pooled to minimize inter-individual variability and provide sufficient input material. Pooled plasma samples were then processed in parallel by SEC and cDLA workflows for side-by-side comparison.

**Data analysis**
Flow cytometry data were analyzed using FlowJo™ Software (Becton, Dickinson and Company). Beads were first identified by gating on forward scatter, side scatter, and bead-specific fluorescence, with singlets further gated using forward scatter parameters. Probe fluorescence intensities were quantified for each bead population as previously described[3] or directly in FlowJo. The fraction of "on" beads was converted to the average number of analyte molecules bound per bead using Poisson statistics and then mapped to analyte concentration using a four-parameter logistic (4PL) calibration curve. All statistical analyses were performed in GraphPad Prism 10, and figures were prepared in GraphPad Prism 10 and Adobe Illustrator.

#### Data availability
The data supporting the findings of this study are available within the paper and its Extended Data files.


#### Acknowledgements
The authors thank Drs. Brian J. Wainger and Christine Marques (Massachusetts General Hospital) for providing the Alzheimer's patient and neurological control samples used in this study. We also thank Mr. Arvin Dayao and Dr. Bogdan Budnik for assistance with Simoa and proteomic experiments, respectively. This work was supported by funding from Good Ventures (to D.R.W.). Y.K. acknowledges NIH grants DP1GM149751 and R01NS112139-01A1 and HFSP grant RGP0032/2022. S.J.Z. is supported by a postdoctoral fellowship (F32 AG087642) from the National Institutes of Health.


#### Author contributions
S.J.Z. conceived the study and designed the experiments. S.J.Z., S.-C.W., and M.L. performed the experiments. S.-C.W. analyzed the micrograph co-localization data. S.J.Z. and Z.J. analyzed the remaining data, including the initial mathematical estimation of experimental parameters. R.F. and Y.K. contributed the lipid–DNA conjugates. S.J.Z. and D.R.W. wrote the manuscript with input from all authors. D.R.W. supervised the study and provided funding.

#### Competing interests
D. R. W. is a founder, equity holder, and Board of Directors member of Quanterix Corporation, which commercializes the Simoa technology. D. R. W.'s interests were reviewed and are managed by Brigham and Women's Hospital and Mass General Brigham in accordance with their conflict-of-interest policies. D. R. W. and S. J. Z. report a provisional patent application (63/708,136) pending and a patent application related to this work pending. All other authors declare no competing interests.

#### Extended Data

**Extended Data Fig. 1| cDLA capture requires lipid–DNA anchors. a,** Control experiments with rEVs ($10^8$) demonstrate that efficient capture requires lipid–DNA anchors. Streptavidin beads alone or streptavidin beads with dB–DNA lacking a lipid anchor showed minimal recovery, whereas the full cDLA workflow yielded robust capture of CD9-positive EVs. **b,** Application of cDLA to human plasma demonstrates enrichment of endogenous EVs. Minimal non-specific binding was observed with streptavidin beads or streptavidin beads + dB–DNA, while cDLA enabled efficient recovery of canonical EV markers CD9 and Alix. Recovery efficiency was calculated as percentage E/(W+S+E) x100%, where E, W, and S represent signal in eluates, washes, and supernatants, respectively. Data represent mean ± standard deviation of duplicate measurements.

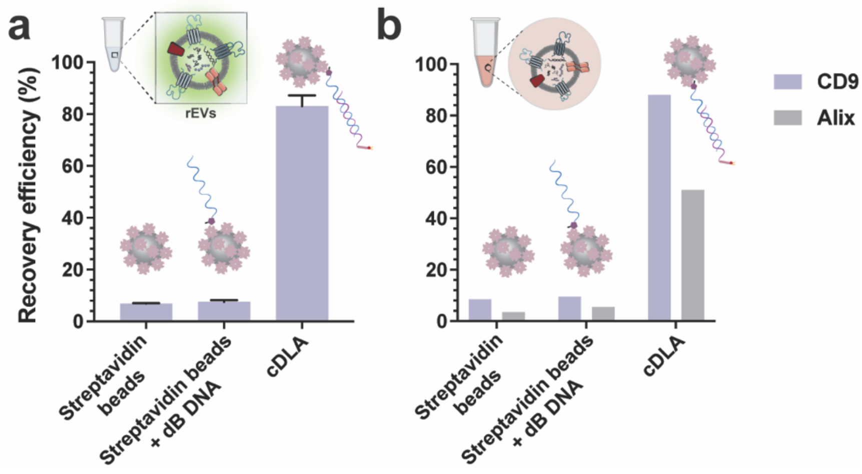


**Extended Data Fig. 2| Optimization of cDLA capture conditions in human plasma.** Human plasma was subjected to cDLA capture under varying conditions. **a,** Effect of lipid–DNA anchor concentration. **b,** Effect of total bead number. **c,** Effect of lipid identity. Recovery efficiency was assessed for CD9 and Alix and expressed as the percentage of signal detected in eluates relative to the total signal across washes, supernatants, and eluates, [E/(W+S+E)] x 100%. Data represent mean ± standard deviation of duplicate measurements.

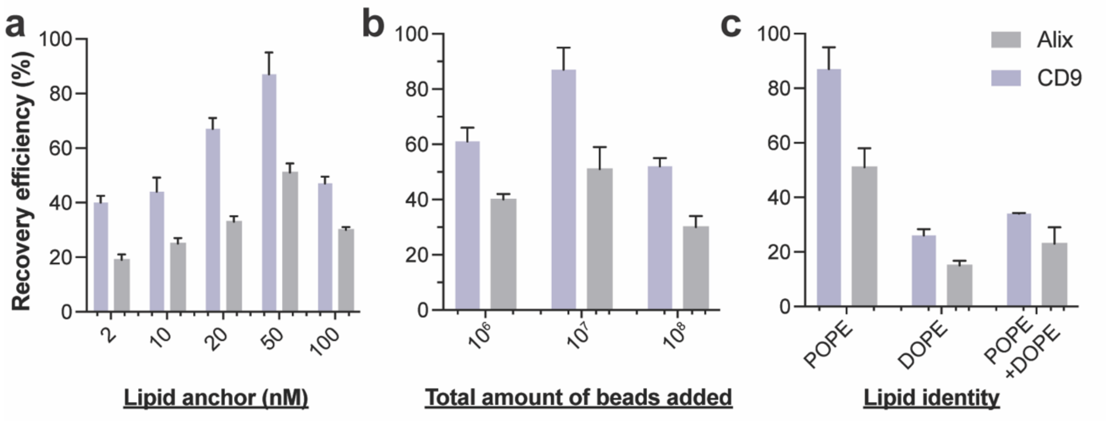

#### Supplementary Figures

**Fig. S1| Comparison of lipid–DNA insertion workflows for EV capture. a,** *Pre-insertion workflow.* DNA–lipid conjugates were first annealed with dB complementary DNA strands to form hybrids, which were then inserted into rEV membranes for 60 min (Step 1), followed by capture on streptavidin-coated beads for 60 min (Step 2). **b,** On-bead insertion workflow. DNA–lipid/dB–DNA hybrids were first immobilized on streptavidin beads for 60 min (Step 1), followed by incubation with rEVs for 60 min to allow on-bead membrane insertion (Step 2). (Right) Recovery efficiency of EV markers CD9 and CD81 under the two workflows, expressed as the percentage of signal detected in eluates relative to the total signal across washes, supernatants, and eluates [E/(W+S+E)] x 100%. Data represent mean ± standard deviation of duplicate measurements.

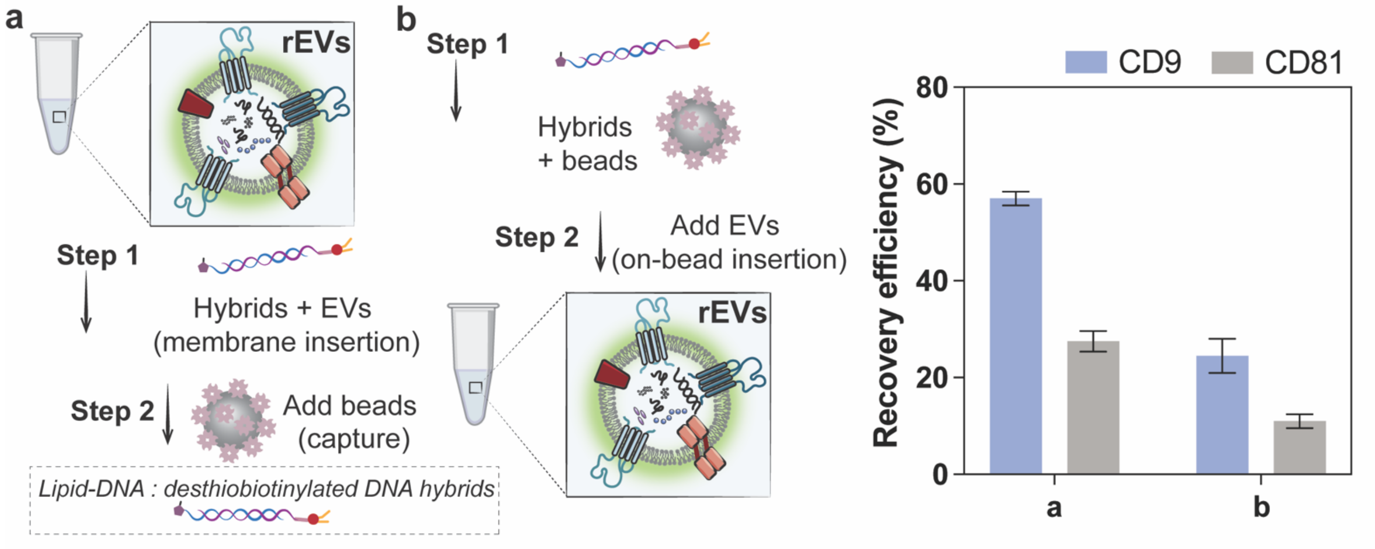

**Fig. S2| Colocalization analysis of GFP⁺ EVs with 750 nm beads.** Probability of colocalization between GFP⁺ rEVs and 750 nm dye-encoded beads across increasing distance thresholds (x-axis). Orange traces show measured colocalization of GFP⁺ EVs with beads, while green traces show colocalization after bead position randomization, serving as a negative control. Data are plotted as probability per bead (y-axis), with mean ± standard deviation shown (n = 10 independent fields of view). Colocalization of GFP⁺ EVs with beads increased with threshold distance, whereas randomized controls remained at baseline, confirming specific bead–EV interactions.

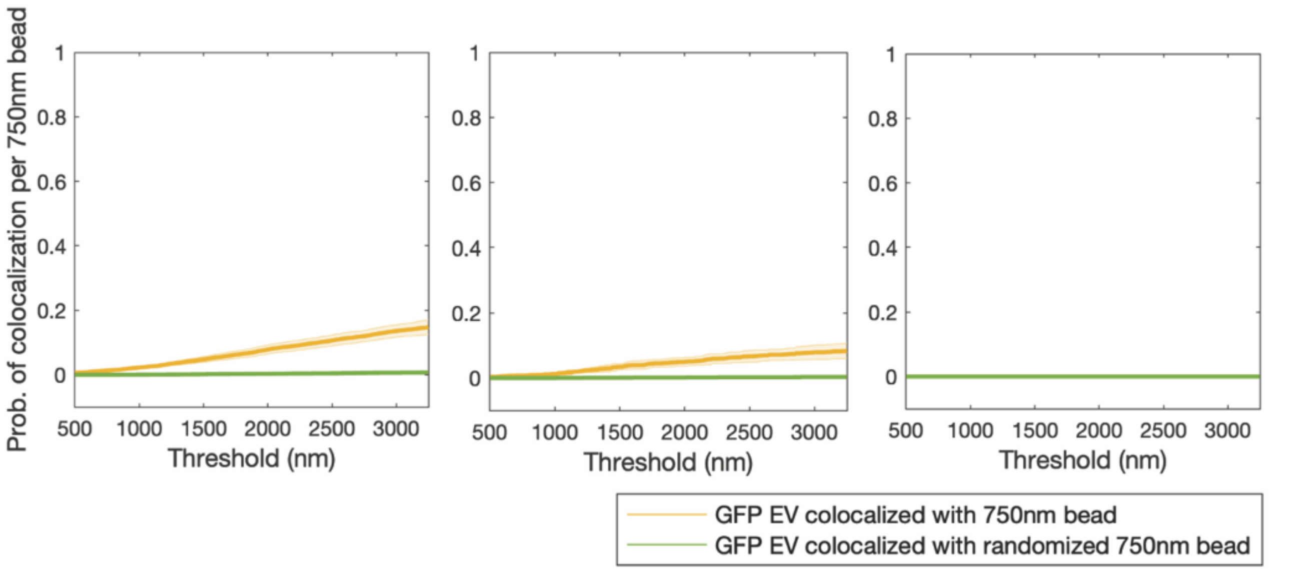

**Fig. S3| Protease protection assay confirms integrity and purity of EVs isolated by cDLA. a,** Schematic of the protease protection assay. NT preserves both internal and external proteins. PK digestion removes external proteins but spares luminal cargo, while permeabilization with PKTx exposes internal proteins to PK digestion. **b,** Quantification of the luminal EV marker Alix (left) and the plasma contaminant Alb (right) in EVs isolated by SEC or cDLA. Alix was protected from PK digestion and lost after PKTx treatment, confirming encapsulation within vesicles. In contrast, Alb was removed by PK alone, consistent with its external association. EVs isolated by cDLA contained markedly higher levels of vesicular Alix and reduced levels of Alb compared to SEC. Data represent three independent trials.

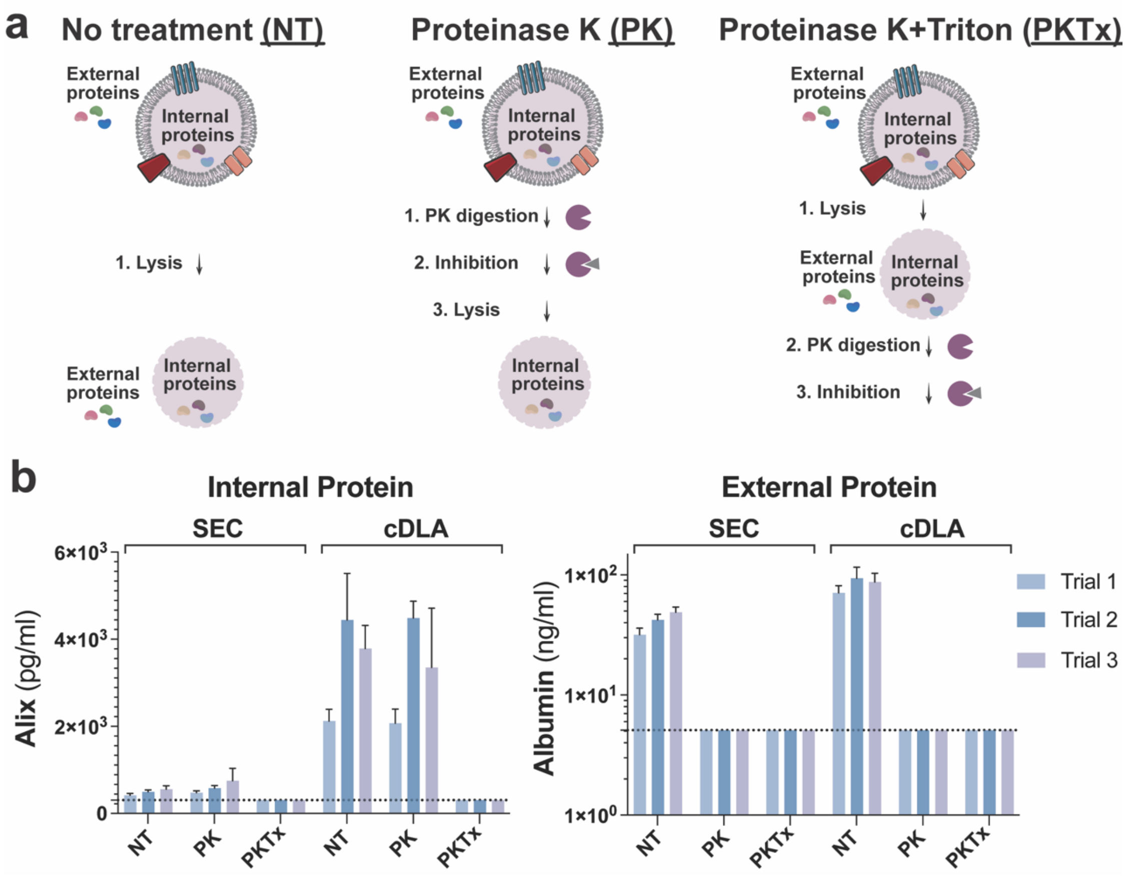

**Fig. S4| Representative LC–MS characterization of lipid–DNA conjugates.** Mass spectra are shown for (a) DSPE–$PEG_8$–DNA, (b) DSPE–PEG600–DNA, (c) DOPE–PEG2000–DNA, and (d) DSPE–PEG2000–DNA. Each spectrum displays the major ion peaks corresponding to the conjugated products. The accompanying table summarizes the calculated (expected) and experimentally determined (observed) molecular masses for each conjugate, confirming successful DNA–lipid coupling through PEG linkers of different lengths.

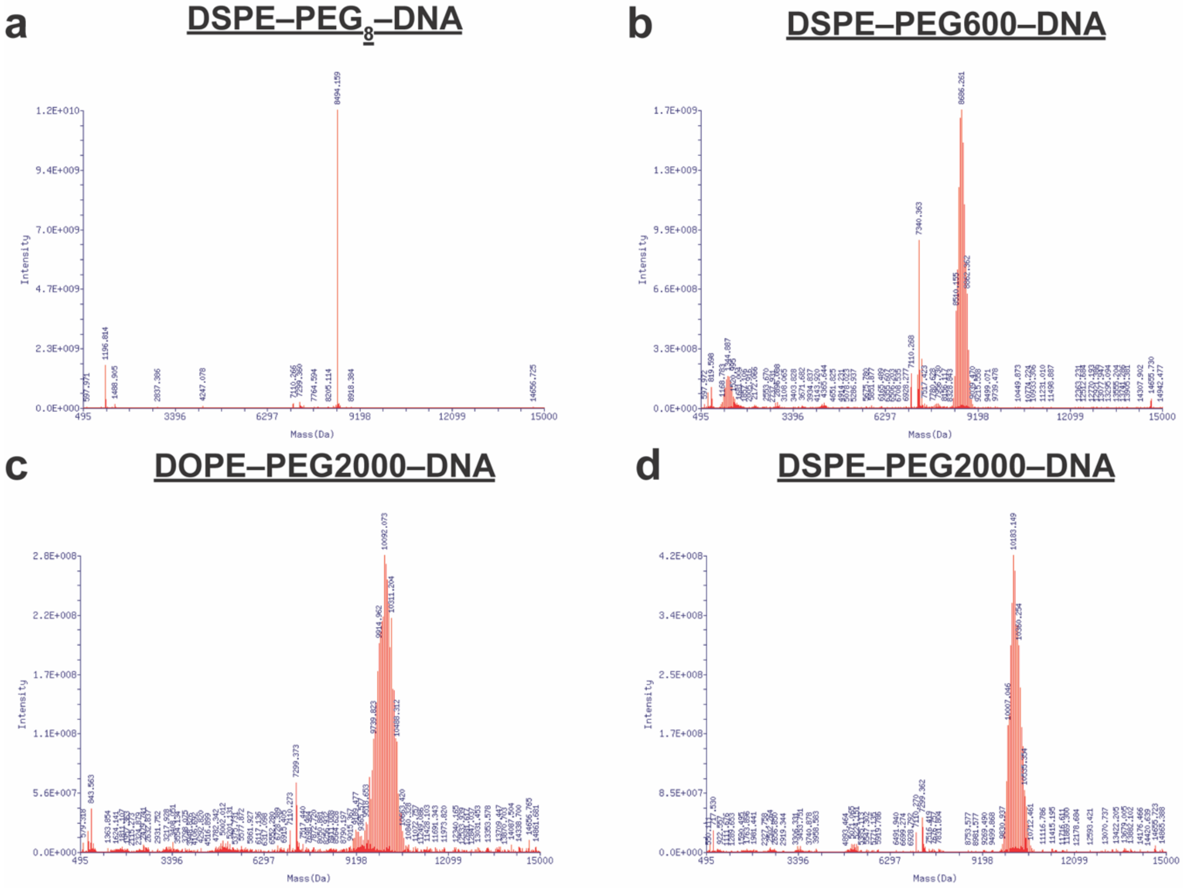


| Compound | Expected Mass | Observed Mass |
|---|---|---|
| a. DSPE-$PEG_8$-DNA | 8498.6 | 8494.159 |
| b. DSPE-$PEG_N$-DNA (MW 600) | 8691 (average) | 8686.261 |
| c. DOPE-$PEG_N$-DNA (MW 2,000) | 10114 (average) | 10092.073 |
| d. DSPE-$PEG_N$-DNA (MW 2,000) | 10115.78 (average) | 10183.149 |

**Fig. S5| Calibration curves for bead-based assays of CD81, CD9, Alix, ApoB, Alb, and tau measured across multiple experimental days.** Data are plotted as AMB versus analyte concentration. Each curve represents an independent calibration replicate performed on different days, demonstrating reproducibility of assay performance. LOD and LLOQ for each analyte are summarized in **Table S7**.

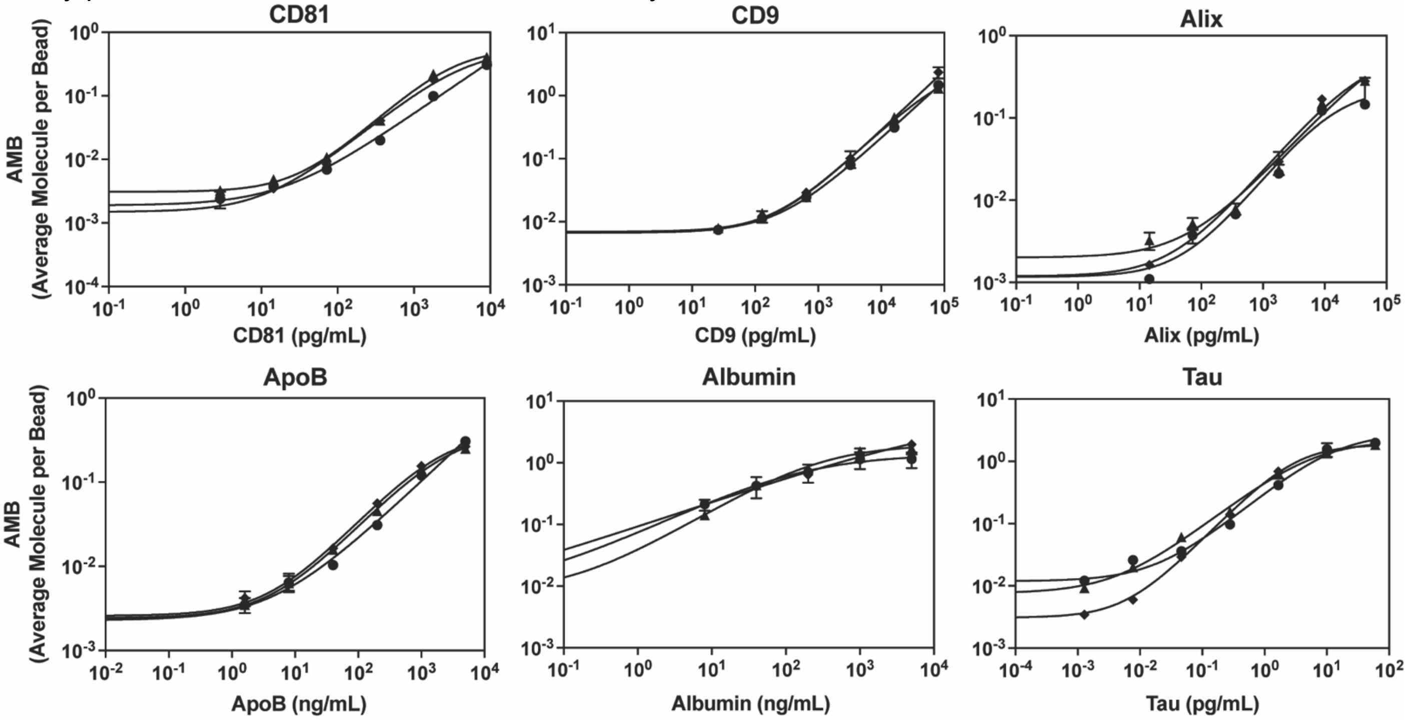

#### Supplementary Tables

**Table S1. DNA sequences for lipid conjugation and complementary oligonucleotides for streptavidin bead capture, imaging, and flow cytometry.** All sequences are listed 5′→3′. Chemical modifications are denoted as follows: /5CarboxyC10/, 5′ carboxyl-C10 linker; /5DBCOTEG/, 5′ dibenzocyclooctyne–triethylene glycol; /5AmMC12/, 5′ amino-C12 modification; /5deSBioTEG/, 5′ biotin–triethylene glycol; /5ATTO647NN/, 5′ ATTO647 fluorophore.

| Application | Sequence (5′→3′) |
|---|---|
| POPE conjugation | /5CarboxyC10/GAG TGC TTT ACA CGG CCG ATG GAG |
| Complementary strand for streptavidin bead coupling | /5deSBioTEG/CTC CAT CGG CCG TGT AAA GCA CTC |
| Complementary strand for testing lipid insertion into EVs (flow cytometry) | /5ATTO647NN/CTC CAT CGG CCG TGT AAA GCA CTC |
| Other lipids (e.g., DSPE, DOPE) conjugation | /5DBCOTEG/TAT ATA TAG GAT CTT GCT GTC T |
| Complementary strand for bead coupling (imaging with color-encoded beads) | /5AmMC12/AGA CAG CAA GAT CCT ATA TATA |
| Complementary strand for testing lipid insertion into EVs (flow cytometry) | /5ATTO647NN/AGA CAG CAA GAT CCT ATA TATA |
| Complementary strand for streptavidin bead coupling | /5deSBioTEG/ACA GCA AGA TCC TAT ATA TA |

**Table S2. Antibodies and recombinant protein standards used in the non-barcoded and barcoded MOSAIC assays. These were selected based on prior studies.[4-6]**

| Analyte | Capture Antibody | Detector Antibody | Recombinant Protein |
|---|---|---|---|
| **CD9** | ab263024 (Abcam) | ab58989 (Abcam) | ab152262 (Abcam) |
| **CD81** | ab79559 (Abcam) | 349502 (Biolegend) | TP317508 (OriGene) |
| **Alb** | DuoSet DY1455 (R&D Systems) | DuoSet DY1455 (R&D Systems) | ab201876 (Abcam) |
| **ApoB** | MAB4124 (R&D Systems) | MAB41242 (R&D Systems) | BA1030 (OriGene) |
| **Alix** | ab275387 (Abcam) | ab117600 Abcam | TP303735 (OriGene) |
| **Tau** | 806501 (Biolegend) | ab174727 (Abcam) | T08-50FN (Sino Biological) |

**Table S3. Sequences of the DNA primer-template, and probe utilized in the barcoded MOSAIC assays in this study.** Bolded sequences denote complementary regions in the primer and template. Asterisks (*) indicate the presence of phosphorothioated nucleic acid bases, incorporated to inhibit exonuclease degradation. The term "invdT" signifies an inverted dT at the 3′-end of the probes, creating a 3′-3′ linkage that prevents degradation by 3′ exonucleases and hinders extension by DNA polymerases.

| | |
|---|---|
| Primer P6 | 5′-Azide TTTTTTTTTTTTTTT TAGACACCGTTCCTTGGACAGA*G*C |
| Primer P13 | 5′-Azide-TTTTTTTTTTTTTTT TAAACGCTTTGAACCGTTAGTT*C*T |
| Template 14 (Pr1) | 5′-phos-ACGGTGTC TATTATGTCCTATCCTCAGC TATTATGTCCTATCCTCA GC TCTGTCCAAGGA |
| Template 15 (Pr12) | 5′-phos-TCAAAGCGTT TA CGAAACAATGGAGATGAG TACGAAACAATGGAGATG AG AACTAACGGT |
| Template 20 (4:1 Pr20:Pr21) | 5′-phos-GAACGGTGTC TACGAGGCAATGG TACGAGGCAATGG TACGAGGCAATGG TACGAGGCAATGG CCTATACTCAGGC TCTGTCCAAG |

| | |
|---|---|
| Probe 1 (Pr1) | 5′-ATTO565-TATTATGTCCTATCCTCAGC - InvdT |
| Probe 12 (Pr12) | 5′-ATTO647-TACGAAACAATGGAGATGAG - InvdT |
| Probe 20 (Pr20) | 5′-ATTO565-TACGAGGCAATGG - InvdT |
| Probe 21 (Pr21) | 5′-ATTO647-CCTATACTCAGGC - InvdT |

**Table S4. Coupling conditions for the antibody-coated capture beads used in this work.** The parameters were selected based on prior studies.[4-6]

| Analyte | Bead Type | Bead Vendor | Starting Bead Number | Coupling Temperature | EDC (µL) | Capture Antibody (µg) |
|---|---|---|---|---|---|---|
| **CD9** | COMPEL™ COOH-modified Dragon Green beads, 3.2 µm | Bangs Laboratories | $4.2 \times 10^8$ | 4°C | 5 | 80 |
| **CD81** | COMPEL™ COOH-modified Glacial Blue beads, 3.2 µm | | $4 \times 10^8$ | 4°C | 5 | 80 |
| **Alb** | 488 multiplex beads | Quanterix | $2.8 \times 10^8$ | RT | 5 | 60 |
| **ApoB** | COMPEL™ COOH-modified Glacial Blue beads, 3.2 µm | Bangs Laboratories | $4.2 \times 10^8$ | RT | 5 | 80 |
| **Alix** | 488 multiplex beads | Quanterix | $2.5 \times 10^8$ | 4°C | 9 | 60 |
| **Tau** | COMPEL™ COOH-modified Glacial Blue beads, 3.2 µm | Bangs Laboratories | $4.2 \times 10^8$ | 4°C | 5 | 80 |

**Table S5. Assay conditions used for the MOSAIC assays in this work.** MOSAIC assays were performed using either a 2- or 3-step format. The listed incubation times include: (1) binding of the target analyte to beads, (2) binding of the detector to the bead–protein complex, (3) the additional step (time in parentheses) in the non-barcoded assay for applying streptavidin–DNA labeling reagents, and (4) the incubation for the RCA reaction. Parameters were selected based on prior studies.[3, 5-7]

| **Barcoded MOSAIC** | | | |
|---|---|---|---|
| **Analyte** | **Assay Bead Number** | **Detector Antibody-DNA (µg/mL)** | **Incubation Times (min)** |
| **CD9** | 20,000 | 0.3 | 60-10-120 |
| **CD81** | 20,000 | 0.45 | |
| **ApoB** | 20,000 | 0.2 | |
| **Tau** | 20,000 | 0.2 | |

| **Non-barcoded MOSAIC** | | | | |
|---|---|---|---|---|
| **Analyte** | **Assay Bead Number** | **Detector Antibody (µg/mL)** | **Streptavidin-DNA (pM)** | **Incubation Times (min)** |
| **Alb** | 20,000 | 0.02 | 100 | 60-10-(10)-120 |
| **Alix** | 25,000 | 3.6 | 250 | 60-10-120 |

**Table S6. Recovery of recombinant proteins spiked into human plasma.** Dilution factors were selected based on previously published studies.[4, 6] Recoveries are reported as mean ± standard deviation from duplicate measurements. Acceptable recovery was defined as 70–130% of the spiked concentration. Results are shown for both pooled plasma and DNase buffer with biotin.

| Spiked protein | Plasma dilution factor | Recovery in pooled plasma (%) | Recovery in DNase buffer + biotin (%) |
|---|---|---|---|
| **CD9** | | | |

| | | | |
|---|---|---|---|
| 16200 pg/mL | 60 | 121 ± 8% | 78 ± 6% |
| 648 pg/mL | 60 | 114% ± 7% | 93 ± 8% |
| **CD81** | | | |
| 1800 pg/mL | 60 | 87 ± 8% | 85 ± 2% |
| 72 pg/mL | 60 | 75 ± 6% | 123 ± 8% |
| **Alix** | | | |
| 900 pg/mL | 6 | 78 ± 6% | 95 ± 8% |
| 180 pg/mL | 6 | 92 ± 5% | 107 ± 4% |
| **Tau** | | | |
| 1.67 pg/mL | 4 | 107 ± 5% | 76 ± 3% |
| 0.05 pg/mL | 4 | 94 ± 8% | 84 ± 11% |
| **Alb** | | | |
| 200 ng/mL | 10,000 | 88 ± 4% | 90 ± 2% |
| 8 ng/mL | 10,000 | 99 ± 3% | 77 ± 11% |
| **ApoB** | | | |
| 200 ng/mL | 500 | 101 ± 9% | 84 ± 9% |
| 8 ng/mL | 500 | 91 ± 7% | 76 ± 6% |

**Table S7. Analytical sensitivities of assays shown in Fig. S5.** LOD and LLOQ were determined for each analyte across three independent experimental days. Values are reported as mean ± standard deviation, with the minimum and maximum values observed across the three days shown in brackets. LODs were defined as three standard deviations above the blank, and LLOQs as ten standard deviations above the blank. Units are shown as pg/mL or ng/mL depending on the analyte.

| Analyte | LOD | LLOQ |
|---|---|---|
| | (pg/mL) | (pg/mL) |
| **CD9** | 16.1 ± 12.7<br>[3.1 – 25.8] | 67.0 ± 35.2<br>[44.1 – 107.5] |
| **CD81** | 7.8 ± 4.9<br>[4.4 – 13.4] | 30.3 ± 10.1<br>[20.6 – 40.8] |
| **Alix** | 11.8 ± 6.2<br>[6.6 – 18.6] | 58.3 ± 26.3<br>[28.7 – 78.8] |
| **Tau** | 0.002 ± 0.002<br>[0.0006 – 0.004] | 0.01 ± 0.01<br>[0.002 – 0.02] |
| | **(ng/mL)** | **(ng/mL)** |
| **Alb** | 0.008 ± 0.004<br>[0.004 – 0.013] | 0.031 ± 0.005<br>[0.027 – 0.036] |
| **ApoB** | 0.3 ± 0.1<br>[0.1 – 0.3] | 7.6 ± 5.9<br>[2.3 – 13.9] |

#### Supplementary Texts

##### A. Lipid identity governs probe insertion and EV recovery

Systematic evaluation of lipid anchors using recombinant EVs (rEVs) revealed that chemical identity is a key determinant of recovery efficiency (**Fig. 1a, iii**). DSPE-based probes showed limited insertion, consistent with the high gel–liquid crystalline transition temperature (~74 °C) of saturated C18 chains. Addition of long PEG spacers (e.g., DSPE–$PEG_{45}$) further impaired recovery, likely by favoring aqueous solubility over membrane partitioning. By contrast, POPE—with unsaturated acyl chains and a flexible ethanolamine headgroup—achieved the highest recovery, highlighting the importance of tail fluidity and headgroup chemistry. Short PEG variants (DSPE–$PEG_{8/10}$) supported intermediate performance, underscoring the tradeoff between solubility and anchoring.

Because these comparisons were performed with rEVs, we cannot directly assess sample purity. However, prior studies have documented that long PEG chains persist as polymeric contaminants in proteomics workflows and can also promote nonspecific adsorption of abundant plasma proteins such as albumin and ApoB, thereby reducing effective purity of EV preparations.[8, 9] These observations provide a mechanistic explanation for why DSPE–PEG anchors, though sufficient for nucleic acid analyses in earlier reports,[10, 11] are poorly suited for proteomics. By contrast, POPE and short-PEG anchors balance insertion efficiency with reduced contamination risk, making them more compatible with downstream proteomic applications.

Together, these results establish general design rules in which lipid tail rigidity, PEG length, and headgroup chemistry jointly dictate insertion and stability, while literature precedent supports their implications for purity. This framework explains the limitations of earlier DSPE–PEG nanoprobes and identifies POPE and short-PEG anchors as superior chemistries for scalable, proteomics-compatible EV isolation.

##### B. Estimation of beads required for capturing EVs

To evaluate whether the bead inputs used in our capture assays were sufficient to accommodate the intended number of EVs, we estimated the bead numbers required to capture $10^8$ rEVs.[12] Beads with diameters of 0.9 µm or 2.7 µm were considered, while rEV diameters were set at 30–150 nm for the calculation—a range commonly reported for exosomes (a subtype of EVs)[13]—which we highlighted as the shaded region in **Fig. S6** to provide a concrete example, although EVs more broadly encompass a wider distribution of sizes and subtypes. Both beads and EVs were approximated as spheres. To estimate steric occupancy on bead surfaces, we used the projected cross-sectional area of EVs as a proxy for their footprint upon binding, rather than the full vesicle surface area. The surface area of a bead ($A_{bead}$) was calculated as $4\pi r^2$, where $r$ is the bead radius, yielding 2.54 µm$^2$ for 0.9 µm beads (r = 0.45 µm) and 22.9 µm$^2$ for 2.7 µm beads (r = 1.35 µm). The projected cross-sectional area of an EV ($A_{EV}$) was estimated as $\pi r_{EV}^2$, giving values of 0.00071 µm$^2$ for 30 nm EVs (r = 15 nm) and 0.018 µm$^2$ for 150 nm EVs (r = 75 nm). From these values, the number of EVs that could theoretically be accommodated on each bead surface was estimated by dividing $A_{bead}$ by $A_{EV}$. For 0.9 µm beads, this calculation predicts between ~30,000 EVs per bead (for 30 nm EVs) and ~1,300 EVs per bead (for 150 nm EVs). Using these bead capacities, the number of beads required to capture all $10^8$ EVs was then calculated. For 0.9 µm beads, between $2.8 \times 10^4$ and $6.9 \times 10^5$ beads are needed depending on EV size, whereas for 2.7 µm beads, the requirement is lower, ranging from $3.1 \times 10^3$ to $7.7 \times 10^4$ beads. This simulation was performed to provide a theoretical framework for bead-to-EV ratios and to justify the starting bead numbers selected in our experimental assays. In addition, this analysis can be generalized to scale capture reactions for different bead sizes, EV populations, or other lipid bilayer vesicles.

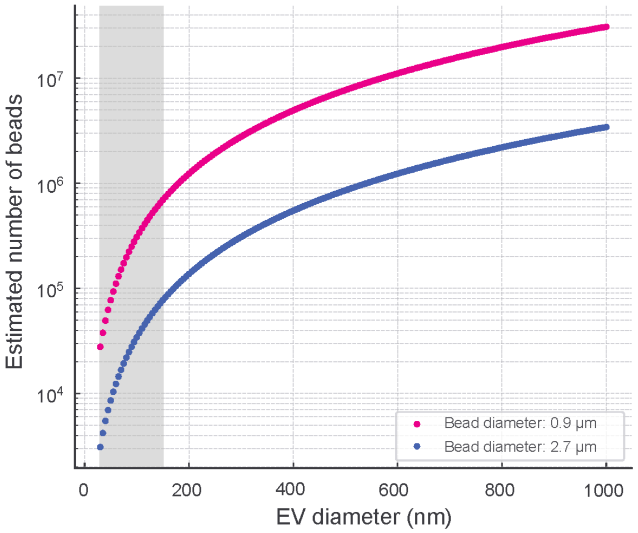


**Fig. S6| Estimated Number of Beads Required for Capturing EVs.** This figure shows the number of 0.9 µm (pink) and 2.7 µm (blue) beads required to capture $10^8$ EVs across a size range of 30 nm to 1000 nm. The shaded region highlights the typical rEV size range. The bead requirements were calculated based on the surface area of the beads and the cross-sectional area of rEVs, assuming spherical geometry and using EV cross-sectional area as a steric footprint.

C. Estimation of lipid probe required for EV capture

To determine the appropriate amount of lipid probe required for lipid-based EV capture, we defined theoretical lower and upper bounds.

*1) Lower bound: one lipid probe per EV.*

The minimal case assumes that each EV incorporates a single lipid probe. For a given number of EVs, $N_{EV}$, the lipid requirement is simply: $N_{lipid}$ = $N_{EV}$. For $N_{EV}$=$10^8$, this corresponds to $10^8$ lipid molecules, or approximately 1.7 x $10^{-4}$ pmol. This value represents a theoretical minimum that is not practically attainable, as EV membranes are heterogeneous and lipid probe incorporation is governed by equilibrium partitioning, with only a fraction of DNA–lipid probes in solution successfully inserting into EV membranes.

*2) Upper bound: bead surface area–based packing density.*

The maximal case assumes complete bead surface coverage by lipid probes. Each phospholipid occupies an area of approximately 0.65 nm$^2$,[14] corresponding to a packing density ($\sigma_{lipid}$) of approximately 1.5 x $10^6$ lipid molecules/µm$^2$. For spherical beads of diameter $d_{bead}$, the surface area is calculated as $A_{bead} = 4\pi\left(\frac{d_{bead}}{2}\right)^2$. The total surface area of $N_{bead}$ is $A_{total} = N_{bead} \times A_{bead}$. The total number of lipid probes required $N_{lipid}$ is then $N_{lipid} = A_{total} \times \sigma_{lipid}$. For 2.7 µm beads, $A_{bead}$=22.9 µm$^2$, giving approximately 3.5 x $10^7$ lipid molecules per bead. If $10^4$ beads are used in a capture reaction, the lipid requirement is approximately 3.5 x $10^{11}$ molecules (~0.6 pmol). For the maximal bead input required to capture $10^8$ EVs (7.7 x $10^4$ beads), the lipid requirement rises to ~ 4.5 pmol. This upper bound likely overestimates the true demand, as DNA–lipid conjugates are bulkier than native phospholipids and steric hindrance further reduces achievable packing densities. These calculations define a lower bound of 1.7 x $10^{-4}$ pmol (one probe per EV) and an upper bound of ~ 5 pmol (complete bead surface coverage under maximal bead usage).

D. Timescale estimate for lipid insertion and EV capture

To determine suitable incubation times and mixing conditions a priori, we estimated the characteristic timescales for (i) insertion of DNA–lipid probes into EVs and (ii) subsequent capture of lipid-modified EVs onto streptavidin-coated beads. These estimates were derived from diffusion-limited theory and prior literature,[15-17] and were used to guide the experimental protocol.

*Stage 1 — Lipid–EV insertion.*

The insertion of amphiphilic DNA–lipid conjugates into EV membranes is expected to occur rapidly. Partitioning of small amphiphilic probes, including carbocyanine dyes, NBD-phospholipids, and PEGylated lipids, into lipid bilayers has been studied extensively and occurs on timescales of 0.1–60 s, depending on lipid tail hydrophobicity and bilayer composition.[18] For DNA–lipid conjugates, the rate-limiting step is diffusion-mediated encounter with vesicles rather than insertion itself, which is energetically favorable and effectively barrierless once contact occurs. Insertion is therefore predicted to be largely complete within minutes, well below the timescales for bead capture (see below). Based on these considerations, we set a conservative insertion window of 30 min at room temperature under 100 rpm orbital mixing. This ensures uniform lipid incorporation even under heterogeneous EV membrane compositions and avoids vesicle aggregation. In practice, extending the incubation to 1 h consistently improved capture efficiency, likely reflecting secondary processes such as diffusion-limited probe–EV encounters in bulk solution, heterogeneous probe loading across vesicle subpopulations, and lateral probe reorganization within membranes that enhances accessibility for bead binding.

*Stage 2 — EV capture on streptavidin beads.*

The slower step is the encounter of lipid-modified EVs with streptavidin-coated beads. Using Smoluchowski diffusion-limited theory, the pseudo–first-order rate constant for capture is: $k' = 4\pi D_{EV} r n_{bead}$, where $D_{EV}$ is the EV diffusion coefficient, $r$ is bead radius, and $n_{bead}$ is bead number density. Applying the Stokes–Einstein relation, the diffusion coefficients for EVs of diameters 30, 100, and 150 nm are ~14.6, 4.4, and 2.9 μm$^2$/s, respectively. For 100 nm EVs ($D_{EV}$ = 4.4 μm$^2$/s) and bead inputs corresponding to 100 μL reactions, the predicted half times were as follows. Using $10^5$ of 0.9 μm beads corresponds to a half-time of approximately 7.7 h. Increasing the bead number to 6.9 x $10^5$ of 0.9 μm beads (upper capacity estimate) results in a half-time of around 1.1h. Scaling with EV size predicts ~3 times faster capture for 30 nm vesicles and ~1.5 times slower for 150 nm vesicles. These values represent theoretical limits under the assumption of perfect sticking efficiency; in practice, steric and orientation constraints reduce effective capture rate. Because pure diffusion predicts multi-hour half-times, we set the bead-capture incubation to 1 h at room temperature, with intermittent orbital shaking at 150 rpm for 10 s every 15 min (0, 15, 30, 45 min). This regimen was chosen prospectively to reduce bead sedimentation and boundary layers, thereby increasing the effective encounter rate relative to diffusion-only predictions. Theoretical estimates suggested that under these conditions, a substantial fraction of EVs would be expected to be captured within 1 h at higher bead loads, with yields approaching ~70% after 2 h.

#### References


1. Saminathan, A. et al. A DNA-based voltmeter for organelles. *Nat Nanotechnol* **16**, 96-103 (2021).
2. Cohen, L. et al. Digital direct detection of microRNAs using single molecule arrays. *Nucleic Acids Res* **45** (2017).
3. Wu, C. et al. High-Throughput, High-Multiplex Digital Protein Detection with Attomolar Sensitivity. *Acs Nano* **16**, 1025-1035 (2022).
4. Norman, M. et al. L1CAM is not associated with extracellular vesicles in human cerebrospinal fluid or plasma. *Nat Methods* **18**, 631-634 (2021).
5. Ter-Ovanesyan, D. et al. Improved isolation of extracellular vesicles by removal of both free proteins and lipoproteins. *Elife* **12** (2023).
6. Norman, M. et al. High-Sensitivity Single Molecule Array Assays for Pathological Isoforms in Parkinson's Disease. *Clin Chem* **68**, 431-440 (2022).
7. Zhang, S.J. et al. A Multiplexed Digital Platform Enables Detection of Attomolar Protein Levels with Minimal Cross-Reactivity. *Acs Nano* **18**, 29891-29901 (2024).
8. Zhang, X.W. et al. Effects of pharmaceutical PEGylation on drug metabolism and its clinical concerns. *Expert Opin Drug Met* **10**, 1691-1702 (2014).
9. Harris, J.M. et al. Effect of pegylation on pharmaceuticals. *Nat Rev Drug Discov* **2**, 214-221 (2003).
10. Wan, Y. et al. Rapid magnetic isolation of extracellular vesicles via lipid-based nanoprobes. *Nat Biomed Eng* **1** (2017).
11. Sun, N. et al. Coupling Lipid Labeling and Click Chemistry Enables Isolation of Extracellular Vesicles for Noninvasive Detection of Oncogenic Gene Alterations. *Adv Sci* **9** (2022).
12. Geeurickx, E. et al. The generation and use of recombinant extracellular vesicles as biological reference material. *Nat Commun* **10** (2019).
13. Chaudhary, P.K. et al. Shedding Light on the Cell Biology of Platelet-Derived Extracellular Vesicles and Their Biomedical Applications. *Life-Basel* **13** (2023).
14. Nagle, J.F. et al. Structure of lipid bilayers. *Bba-Rev Biomembranes* **1469**, 159-195 (2000).
15. Baumgart, T. et al. Fluorescence probe partitioning between Lo/Ld phases in lipid membranes. *Bba-Biomembranes* **1768**, 2182-2194 (2007).
16. Sezgin, E. Plasma membrane labelling efficiency, internalization and partitioning of functionalized fluorescent lipids as a function of lipid structure. *Rsc Chem Biol* (2025).
17. Grebenkov, D.S. Diffusion-Controlled Reactions: An Overview. *Molecules* **28** (2023).
18. Mcintyre, J.C. et al. Fluorescence Assay for Phospholipid Membrane Asymmetry. *Biochemistry-Us* **30**, 11819-11827 (1991).
